# Pose AI prediction of neurological status in the Neuroscience Intensive Care Unit

**DOI:** 10.1101/2025.03.12.25323510

**Authors:** Rui Feng, Florian Richter, Elizabeth Mari, Alec Gleason, Chi Le, Christopher Kellner, Raj Shrivastava, Madeline Fields, Benjamin Rapoport, Joshua Bederson, Eric Es Schadt, Benjamin S. Glicksberg, Felix Richter, Neha Dangayach

## Abstract

**BACKGROUND:** The neurological exam is pivotal in assessing patients with neurological conditions but has severe limitations: it can vary between examiners, it may not discern subtle or subacute changes, and because it is intermittent can delay recognition of new deficits. Even in the Neuroscience Intensive Care Unit (NSICU), neurological exams are conducted only hourly. The majority of ICUs/hospitals lack subspecialized neurocritical care services, exacerbating this neurologic monitoring gap. We hypothesized that pose AI, a machine learning approach to track patient position, could provide a continuous and relevant method of neurological monitoring.

**METHODS:** We retrospectively collected video segments from patients in the NSICU and the Epilepsy Monitoring Unit (EMU) who underwent video-EEG at a large, urban hospital between July, 2024 to January, 2025. We externally validated two leading pose AI models, ViTPose and Meta Sapiens. We then developed a robust movement index and evaluated its correlation with two measures of consciousness obtained through hourly physical exams, the Glasgow Coma Scale (GCS) and Richmond Agitation Sedation Scale (RASS).

**RESULTS:** We collected 998,520 video minutes from 119 NSICU and EMU patients. ViTPose demonstrated superior performance to Sapiens across multiple metrics, so we used ViTPose to calculate a computer vision movement index (λ_MI_). We observed higher movement with increasing GCS (GCS 3–8 λ_MI_=0.52, GCS 9–13 λ_MI_=0.70, GCS 14 λ_MI_=3.52, GCS 15 λ_MI_=10.99, *P*=0.01), a 21-fold increase from the lowest to highest tranche. We also observed 10-fold higher movement in awake/agitated patients (RASS>-1 λ_MI_=6.59) compared to those who were asleep/sedated (RASS≤-1 λ_MI_=0.67, *P*=0.005). Taken together, we developed a novel computer vision movement index and demonstrated expected correlations with GCS and RASS scores in NSICU patients.

**CONCLUSION:** We show that pose AI can provide minimally invasive, continuous and clinically relevant neuro-monitoring in critically ill patients. Neurological conditions account for the highest global disease burden and pose AI may be a low-cost, explainable, and scalable AI solution to address this pressing need for neuro-telemetry.

## INTRODUCTION

The neurological exam is the most crucial component in assessing patients with neurological conditions and drives decision making for time-sensitive interventions including neurosurgery. This exam suffers from several limitations. It is conducted at arbitrarily set time intervals, which can be delayed relative to the onset of new deficits, has inter-examiner variabilities, and may not discern subtle and/or acute changes ^1^. Neurologically critically ill patients may be admitted to a Neuroscience Intensive Care Unit (NSICU) staffed with specialized providers and nurses or to a mixed medical-surgical intensive care unit (ICU). Even in an NSICU, neurological exams are conducted and documented periodically, typically no more than on an hourly basis. These examinations can become disruptive to normal sleep patterns, which has been shown to worsen ICU delirium and prolong length of stay ^2^. Intermittent examinations therefore can create a gap between the fluctuating neurological status of critically ill patients and the recognition of important changes. At the majority of hospitals and ICUs, this gap in neurologic monitoring is even wider because they are not staffed with subspecialized neurocritical care providers and nurses ^3^. In the US, dedicated NSICUs are concentrated in large urban academic hospitals accessible by less than a quarter of the US population ^4^. Thus, a majority of neurologically critically ill patients are monitored in non-specialized settings.

It has been hypothesized that continuous monitoring of neurosurgical function, neuro-telemetry can provide a solution to the shortcomings of intermittent examination. Sedation monitoring is a salient example of the unmet need for neuro-telemetry. Sedation management is important for all ICUs, and there are over 120,000 ICU beds in the US ^5^. Cost of medications used for sedation has been estimated to be at $27,603 per patient ^6^. A wide range of issues have been associated with over- or under sedation: accidental extubation, accidental device removal, prolonged mechanical ventilation, increased ICU delirium, exacerbation of underlying brain injury, pain and psychological trauma ^7–10^. In neurologically critically ill patients, titration of sedation is even more challenging since it can impede accurate detection of neurological deterioration. The current standard of care for sedation monitoring in the ICU is to use validated instruments that rely on the human-performed neurologic exam such as the RASS, which are inherently non-continuous ^11–13^. Electroencephalography (EEG) is limited in its utility, given that the full montage is cumbersome to apply and maintain uninterrupted signal with frequent adjustments by trained staff required, access to EEG machines and technicians is limited, subspecialized training in epilepsy is needed for formal interpretation, and interpretations are often made on time scales of hours to days after clinically relevant events^14^. The bispectral index (BIS), a processed form of EEG, is used in patients undergoing anesthesia and its adoption in ICUs and the operating room has been shown to address some of EEG’s limitations. However, it has unclear validity in non-paralyzed patients and several limitations in monitoring critically ill patients ^15,16^.

Concurrently, there is a vast amount of patient data collected from videos, such as patient monitoring devices and video electroencephalography (video-EEG). This data is not routinely stored in an accessible manner for researchers to analyze and thus grossly underutilized. Preliminary work with computer vision on patient video data has demonstrated value in seizure detection ^17^, Parkinson’s disease ^18^, and cerebral palsy ^19^. Our group previously demonstrated that computer vision could accurately predict sedation and cerebral dysfunction in infants in the ICU ^20^. However, this technology has been incompletely studied in adult inpatient and critical care settings for neurological exam monitoring.

We hypothesized that pose artificial intelligence (AI), a computer vision machine learning approach to track position of anatomical landmarks, has the potential to provide a continuous and accurate method of neurological monitoring as a form of “neuro-telemetry”. To test this hypothesis, we externally validated two recently available pose AI models and compared their performance in two real-world adult inpatient populations. We then developed a movement index from the pose AI outputs and demonstrated encouraging performance correlating with clinical scores on patients’ state of consciousness on the Glasgow Coma Scale (GCS) and Richmond Agitation Sedation Scale (RASS).

## METHODS

### Study design and data acquisition

We implemented a retrospective observational study design and collected data from July 01, 2024 to January 01, 2025. The Institutional Review Board at Icahn School of Medicine at Mount Sinai approved this study. We used broad inclusion criteria and included all patients admitted to the Neuroscience Intensive Care Unit (NSICU) at the Mount Sinai Hospital who underwent video-EEG monitoring during the study period. We also included patients from the Epilepsy Monitoring Unit (EMU) as a control group, as these patients are inpatient but not critically ill or sedated. We collected video-EEG data from the Natus Database (version 9) and transferred all video data through an encrypted connection directly to Mount Sinai’s HIPAA-compliant supercomputing cluster for storage and analysis. We gathered patient demographics, including self-reported race and ethnicity, and relevant clinical data from Electronic Health Records (EHR). Our dataset contains racially and ethnically diverse patient populations with the intention that our results will be generalizable across race, sex, age, and diagnoses.

### Pose AI model implementation and evaluation

We first aimed to determine if pose AI can accurately model neurological status in the hospitalized patient population. We chose two representative and leading pose AI models, ViTPose ^21^ and Meta AI Sapiens ^22^. Prior to Sapiens, ViTPose was thought to be a state of the art pose AI model for Common Object in COntext and OCHuman datasets ^23,24^, and leverages pre-trained vision transformers on 15 million images to extract features from a given person instance ^21,25^. Sapiens is the newest 2D pose estimation model published by Meta AI in 2024 and pre-trained on over 300 million human images ^22^. These models can convert 2D human form to vectorized pose, which consists of X and Y coordinates for each of the standardized set of seventeen anatomical landmarks (*e*.*g*., nose, left/right ear, left/right shoulder, left/right knee) and a likelihood score for each landmark in each frame. Both models were used in their out-of-box state, as they were designed and shown to have high generalizability across a wide range of human pose tasks. We specifically examined the models in their out-of-box state, rather than fine-tuning these pose AI models to our dataset, for three reasons: (i) to evaluate their out-of-distribution generalizability on two external validation cohorts, (ii) to examine representative real world model performance that is likely to be more consistent and stable across different healthcare scenes due to the model’s uniquely large training dataset size, and (iii) to establish a framework for evaluating future models as they come online. In contrast, fine-tuning may improve performance in the unique ICU setting but requires substantially larger datasets, risks overfitting to our own data, and is currently not feasible in the majority real world healthcare environments, which are not connected with the massive GPU compute clusters required for such fine-tuning.

To evaluate how these two leading computer vision models perform in real-world healthcare scenes, we randomly sampled one frame per patient and labeled up to seventeen landmarks per frame. The same frames were put through ViTPose and Sapiens. We generated a receiver operating characteristics area under the curve (ROC-AUC) based on the model’s ability to correctly predict whether an anatomic landmark was visible using the models’ built-in confidence scores. We then calculated the accuracy, sensitivity, specificity, and percentage of high-confidence predictions for each model in predicting landmark visibility. Lastly, we measured the per-landmark position error, which is the Euclidean distance in pixels between predicted anatomical landmark coordinates and their manually labeled coordinates.

### Movement index calculation

Our methodology of summarizing overall movement was similar to reports using pose AI analysis of infant video data, correlated with sedation and cerebral dysfunction ^20^. This approach mirrors extensive work on accelerometry/actigraphy in ICU patients with wearables, which has consistently found that simple movement indices such as variance are highly predictive of multiple ICU and neurologic outcomes, including GCS and RASS^26–28^. Our proposed movement index, derived from pose AI using video data, overcomes limitations of wearable accelerometry devices including signal artifacts, miscalibration, inter-limb discrepancies, need for monitors on multiple limbs, sensor detachment, pressure injury monitoring, and inability to visualize or replay the context in which neurologic changes occurred^29^. After an interim analysis, we selected the superior performing pose AI model to further analyze the videos and developed a movement index (λ_MI_) reflective of a patient’s neurological status. From the collected video segments, we randomly sampled two-minute clips per segment. We filtered for landmarks with high prediction confidence with a confidence threshold of 0.8 and then for frames that contain at least three landmarks above threshold. We then calculated the variance from the X and Y coordinates of each anatomical landmark in the remaining frames which inherently provides robustness to translation, rotation, and outliers. The only modification to the prior pose AI analysis is the variance is computed through a covariance matrix, which captured diagonal movement (*i*.*e*., in both the X and Y direction) better than the single dimensional variances previously used ^20^. The two variance values were extracted from the covariance matrix along the major axis through standard Gaussian Whitening techniques ^30^. We also normalized the variance to the patient’s nose to mid-shoulder distance in each clip to account for body size and camera distance. We then took the maximum of the normalized variance as the movement index (λ_MI_) for the two-minute interval, similar to previous work ^20^.

### Clinical outcomes

To examine the utility and predictive capability of pose AI on patients’ neurological status, we correlated our movement index (λ_MI_) with the two clinical scales routinely collected by nurses based on bedside neurological examinations, the GCS and the RASS. We chose to use the Glasgow Coma Scale (GCS) because it is one of the most widely used tools for standardized assessment of neurological function. Introduced in a landmark study 50 years ago, the GCS is the sum of three subscales (motor, verbal, eye-opening) and ranges from 3 to 15 ^31^. Before its introduction, assessments of consciousness were inconsistent and unreliable ^32^. The GCS is a simple, objective, and reliable measure, and consequently has become internationally adopted as a cornerstone of neurological evaluation across diverse clinical settings and injury types. One limitation of GCS is inconsistent inter-rater reliability of specific point assignments; however, reliability is excellent when grouped into clinically relevant ranges ^1,33^. We therefore grouped our GCS outcomes into these ranges. Another key tool in intensive care is the Richmond Agitation Sedation Scale (RASS) ^11,12^, which monitors depth of sedation and agitation via a 10-point scale ranging from -5 (unarousable) to +4 (combative) ^13^. It was first published almost 25 years ago and has been widely adopted for its ease of use, excellent inter-rater reliability, construct validity, and inclusion of both agitation and sedation ^13,34,35^. Both GCS and RASS are documented in NSICUs as part of standard of care, have high correlation (r>0.90), and drive clinical decision-making including intubation and neurosurgical device placement ^13^.

Both of these scales are documented at one-hour-intervals in the NSICU, so only the NSICU cohort was used for this analysis, not the EMU’s. GCS and RASS scores temporally closest to the time of video capture were recorded. Continuous sedative and paralytic medications infusing at time of video capture were also collected. We compared the movement index (λ_MI_) over two-minute intervals between typical clinically relevant groupings for GCS and RASS. The GCS groupings were 3–8, 9–13, 14, and 15, which are standard stratifications used to define severity of traumatic brain injury (TBI) with high inter-rater reliability ^33,36,37^. We used ordinal logistic regression to determine if there was an association between GCS tranches and movement index. For RASS, we divided our cohort into groups with RASS ≤ -1, and RASS >-1, the typical target for adequate sedation in the NSICU, and used ordinal logistic regression to determine if there was an association. Statistical significance is set at a *P* value of less than 0.05. We also empirically identified the inflection point in movement index across the entire range of RASS from -5 to +3 using nonlinear least squares fit to a sigmoid function.

## RESULTS

### Patient characteristics

To externally validate the AI pose models, we collected video data from two inpatient cohorts (**Figure 1**). Patients in the NSICU are managed for life-threatening conditions affecting the central nervous system, whereas patients in the EMU are admitted for seizure monitoring but are otherwise stable. From July 01, 2024 to January 01, 2025, there were 88 consecutive patients from the NSICU who underwent video-EEG. There were 552 video segments recorded, totaling 321,785 minutes (median 2,591 minutes/patient). From September 01, 2024 to October 31, 2024, there were 31 patients from the EMU who underwent video-EEG. There were 137 video segments recorded, totaling 84,347 minutes (median 1,067 minutes/patient). The demographic characteristics of our cohort are reflective of the diverse racial and ethnic background population of New York City (**Table 1**).

**Table 1.**
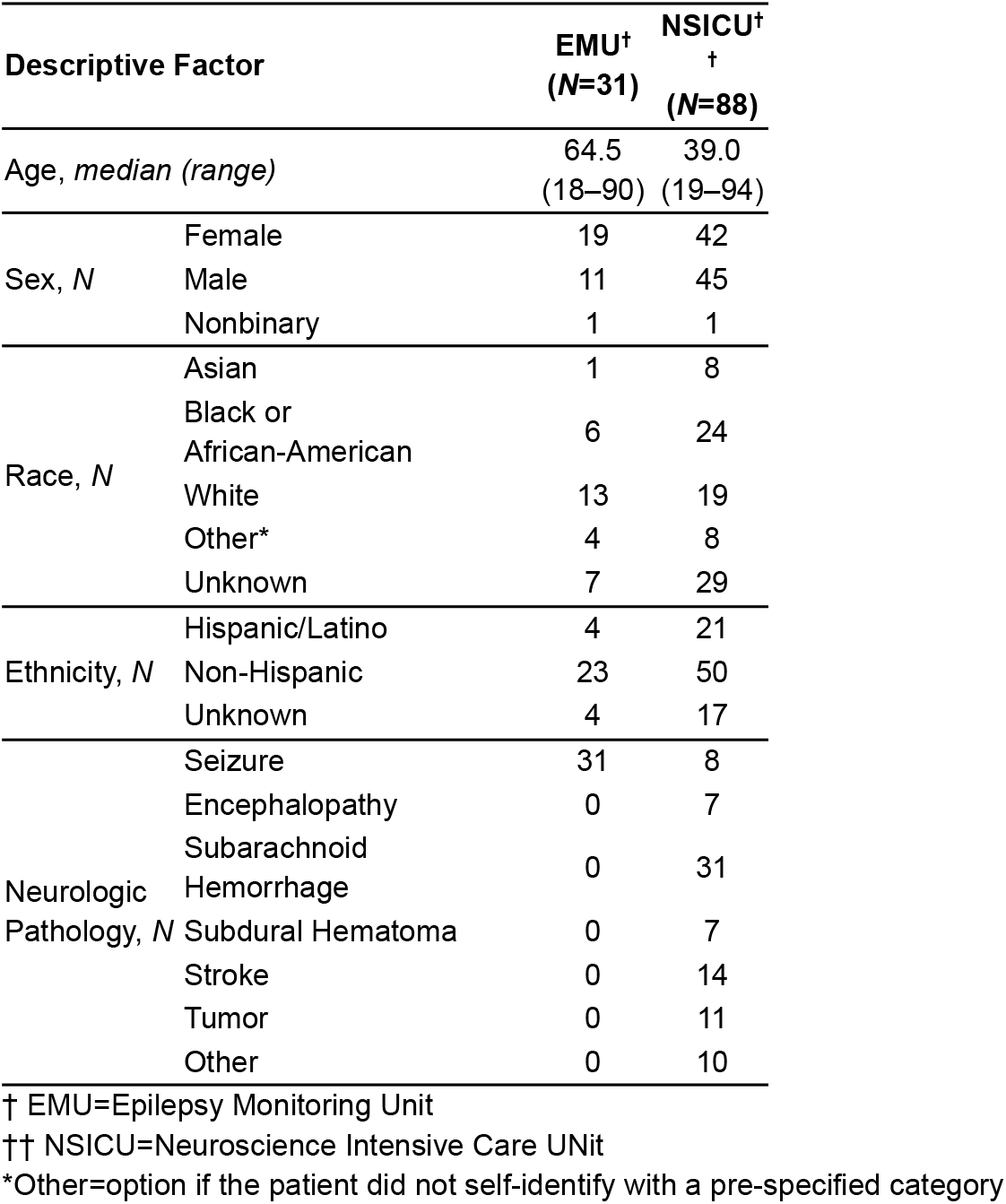
Clinical characteristics of all patients included in this study.

**Figure 1.**
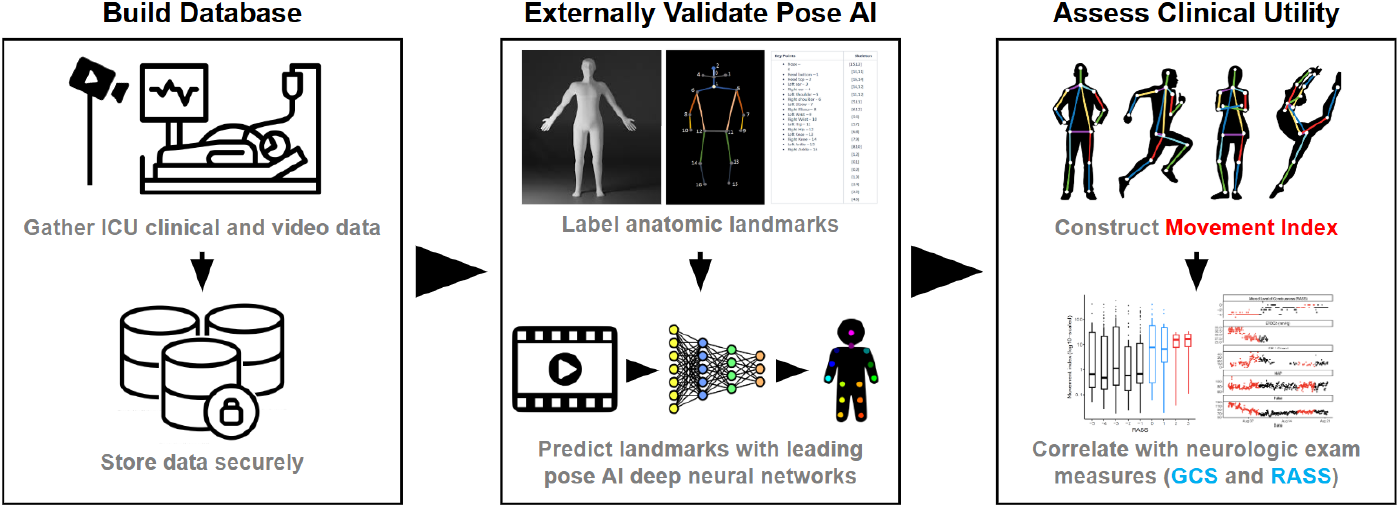
Application of Pose AI to adults requiring inpatient neurologic monitoring. We built a large database of video-EEG data (N=120 adults, 406,132 minutes of video at 30 frames per second) that is stored on a HIPAA-compliant supercomputing cluster ^20^. The dataset comprises two unique inpatient populations that require neurologic monitoring, patients in the epilepsy monitoring unit (EMU) who are being monitored for seizures and patients in the Neuroscience Intensive Care Unit (NSICU) who are being managed for life-threatening conditions affecting the central nervous system. We externally validated two cutting edge pose AI algorithms, ViTPose ^21^ and Meta AI Sapiens ^22^, on gold-standard manually labelled video frames. We then constructed an intuitive movement index derived from pose AI features and demonstrated clinical utility through benchmarking with multiple well-validated neurologic exam instruments.

### Pose AI model performance and external validation

We implemented two leading foundation models in computer vision, ViTPose and Meta AI Sapiens (see **Methods**). These models were originally trained on millions of images to recognize and track human anatomic landmarks. To test their external validity, we randomly sampled one frame per patient from each of the 119 patients, manually labeled each frame, and used standard pose AI metrics to compare model predictions to our ground truth labels. We first evaluated the ability of each model to correctly discern if an anatomic landmark was visible (*i*.*e*., landmark occlusion). ViTPose demonstrated superior performance on the ROC-AUC with 0.74 across both cohorts, compared with Sapiens at 0.68 (**Figure 2a**). Both models performed better on the EMU cohort compared with the NSICU cohort: ViTPose achieved 0.81 for EMU, 0.70 for NSICU, and Sapiens 0.70 for EMU, and 0.67 for NSICU (**Figure 2b**). We then used strict filters to retain only frames with multiple high quality landmark predictions (**Supplemental Feigure 1**). ViTPose had a much higher percentage of frames with high confidence predictions than Sapiens, with high confidence for 57%, compared to Sapiens with only 15%. ViTPose had a median per landmark pixel error of 6.03 pixels and Sapiens was 2.09 pixels, both less than the typical range of human label variability (6.25 pixels, scaled to our video resolution) ^38^. ViTPose also demonstrated superior accuracy and sensitivity, with 0.71 and 0.48 respectively, as compared to Sapiens of 0.61 and 0.11. Sapiens achieved slightly higher specificity at 0.99, while ViTPose was 0.88.

**Figure 2.**
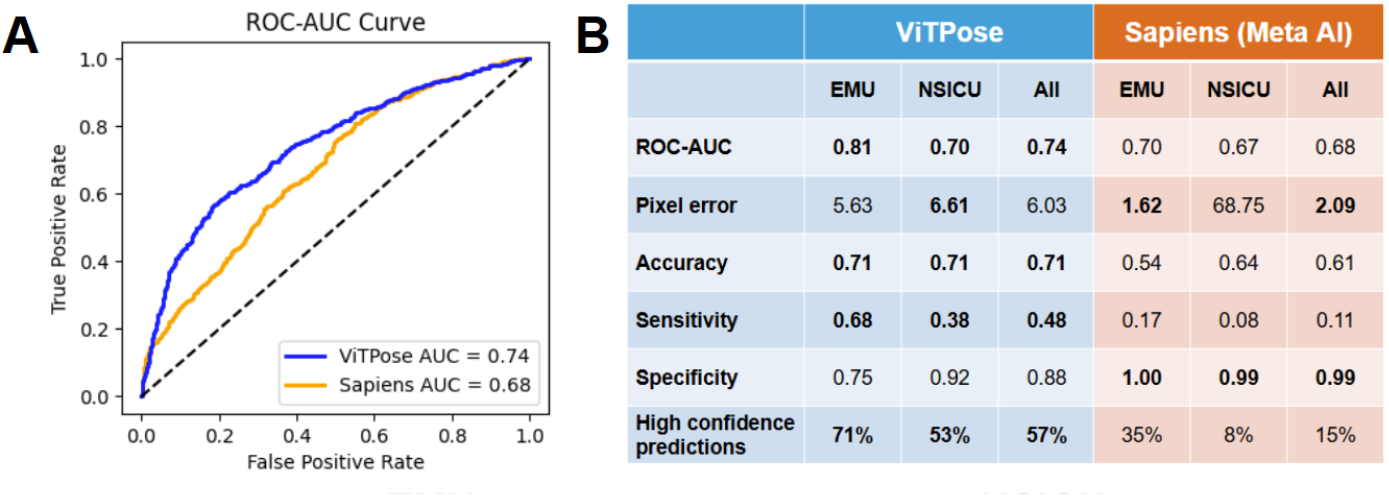
External validation of two leading pose AI models on two inpatient populations requiring neurologic monitoring. **(a)** ViTPose (blue) had a higher receiver operating characteristic area under the curve (ROC-AUC) compared to Sapiens (yellow) when evaluating landmark visibility. **(b)** ViTPose had higher performance compared to Sapiens in almost four times as many healthcare scenes (57% vs 15%) as well as superior ROC-AUCs, accuracy, and sensitivity with slightly lower specificity. Performance was higher in the EMU compared to NSICU across every metric, likely reflecting the lower complexity of the healthcare scene.

Overall, across two external validation cohorts, ViTPose had high performance in almost four times as many healthcare scenes as Sapiens (57% vs 15%) as well as superior ROC-AUCs, accuracy, and sensitivity with slightly lower specificity. In addition, on review of illustrative example frames, ViTPose correctly identified patients and anatomic landmarks where Sapiens did not. We therefore proceeded to use ViTPose for all downstream analyses because it was the model with better generalizability to real world inpatient healthcare scenes.

### ViTPose performance across subgroups

We broke down ViTPose’s performance across all body regions and found it performed best in anatomical landmarks on the head, similar to previous literature ^20,39^ (**Supplemental Table 1**). We observed consistent algorithm performance across multiple demographic subgroups (**Figure 3A**). Specifically, there were no statistically significant differences in ViTPose prediction accuracy between race (Kruskal-Wallis *p=0*.*12*), ethnicity (*p=0*.*19*), self-reported gender (*p=0*.*84*), or age groups (*p=0*.*15*). We summarized the primary neurological diagnoses into seven broad categories and also did not find a difference in prediction accuracy (Kruskal-Wallis *p=0*.*96*, **Figure 3B**).

**Figure 3.**
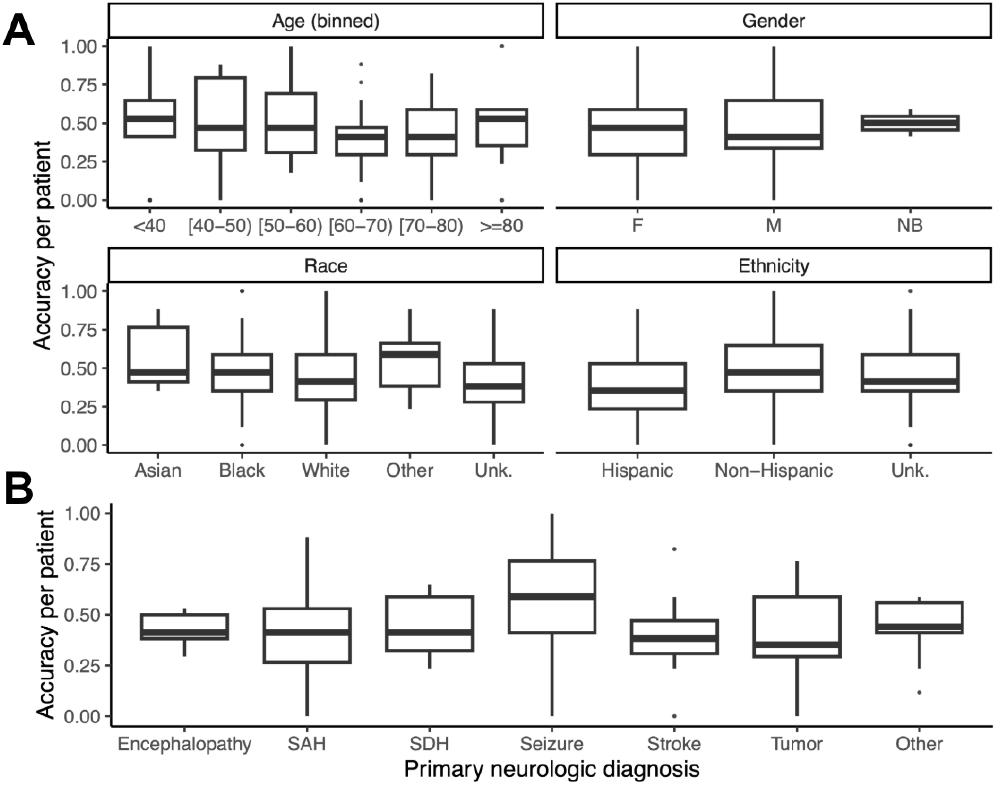
Comparison of pose AI performance across demographic groupings and neurologic diagnoses. **(a)** Accuracy of pose AI (y-axis) was not statistically significantly different within age or self-reported gender, race, and ethnicity (subplots, all four Kruskal-Wallis *P*>0.1). **(b)** Accuracy of pose AI (y-axis) was not different between primary neurologic diagnoses (x-axis, Kruskal-Wallis *P*=0.96). Box plots show the median and 25^th^/75^th^ quartiles; whiskers show the bounds of 1.5 times the interquartile range.

### Association between patient movement, predicted with pose AI, and validated neurological exam measures

We used pose AI features from ViTPose to construct a movement index (λ_MI_, see **Methods**) as a global metric of a patient’s movement. To evaluate the clinical utility of this movement index, we performed association tests with two neurological exam measures that are documented hourly in the NSICU, the GCS and RASS. The GCS and RASS have high correlation in our data (Spearman’s ρ=0.83, *p*=10^−96^), consistent with the literature ^13^ and the notion that they fundamentally measure similar patient clinical state changes (*i*.*e*., movement). We observed higher movement with higher GCS tranches (GCS 3–8 λ_MI_=0.52, GCS 9–13 λ_MI_=0.70, GCS 14 λ_MI_=3.52, GCS 15 λ_MI_=10.99, ordinal logistic regression *P*=0.01), a 21-fold increase between the lowest to highest tranche (**Figure 4a**). The markedly higher patient movement observed at GCS of 14 and 15 are consistent with these patients having typically normal neurologic function, while lower GCS tranches are almost universally observed in patients with abnormal neurologic exams. We also observed higher movement indices with higher RASS scores (**Figure 4b**), with a 26-fold increase between the RASS -5 (λ_MI_=0.67) and RASS +2 (λ_MI_=17.17). Movement was 10-fold higher in awake/agitated patients (RASS>-1 λ_MI_=6.59) compared to those who were asleep/sedated (RASS≤-1 λ_MI_=0.67, ordinal logistic regression *P*=0.005). Furthermore, consistent with RASS between -1 and 0 being the typical target for achieving sufficient sedation based on patient movement, we empirically identified an inflection point in movement index at RASS -0.28 based on the best-fit sigmoid function. Overall, our movement index was statistically significantly associated with both RASS and GCS, with prominent changes in the movement index observed at clinically relevant inflection points.

**Figure 4.**
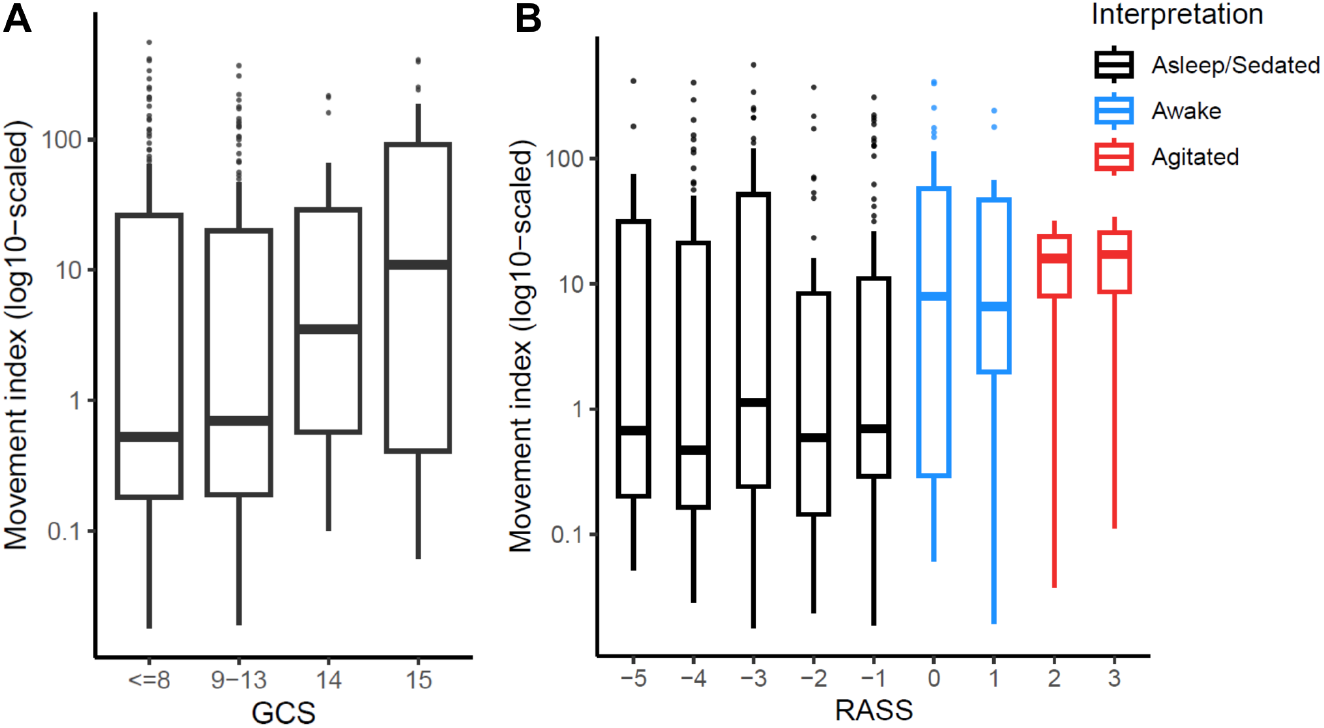
Association between a movement index constructed from ViTPose and two key validated neurologic exam instruments. **(a)** There was a statistically significant increase in movement (y-axis, log-10 scaled) in higher tranches of the Glasgow Coma Scale (GCS, x-axis), with a 21-fold increase between the lowest and highest groupings (ordinal logistic regression *P*=0.01). **(b)** Movement (y-axis) across all Richmond Agitation Sedation Scale (RASS) scores (x-axis) demonstrated a statistically significantly lower movement in asleep/sedated patients (black, ordinal logistic regression *P*=0.005) compared to those who were awake/agitated patients (blue/red). Box plots show the median and 25^th^/75^th^ quartiles; whiskers show the bounds of 1.5 times the interquartile range.

## DISCUSSION

In this study, we found that pose AI can accurately model patient pose in hospital settings including ICU environments and that its output meaningfully correlated with clinical neurological status. Previous studies in computer vision looked at patient videos in the outpatient setting or in a relatively functional inpatient population ^17–19^. The ICU population is unique in that patients are critically ill, commonly sedated, and/or on mechanical ventilation, and consequently require more intensive neuromonitoring. This study represents an important first step toward validating how AI, particularly computer vision of continuous patient video feeds, can be utilized as a tool for improving neurological assessments in a real-world critical care setting.

We chose two state-of-the-art pose AI models, ViTPose and Meta AI Sapiens. ViTPose is a well-established model that has demonstrated robust performance on benchmark testing ^21^. Meta AI Sapiens is the newest computer vision model pre-trained on over 300 million human images and has better performance on pose estimation tasks when compared head-to-head against ViTPose ^22^. We found ViTPose to be more accurate in our tasks than Meta AI Sapiens with a higher proportion of high confidence landmarks. We expect that performance will greatly improve with fine-tuning on datasets with characteristics reflective of our inpatient population. Importantly, we demonstrated that the pose AI predictions did not differ in accuracy across broad age, gender, racial, ethnic, and primary neurological diagnosis subgroups. This indicates its robust generalizability across different population and disease states. In addition, our video data varied significantly in image resolution, camera angle, distance, and patient visibility, which serves as a promising indicator that pose AI can generalize to different healthcare settings with inconsistent video quality.

We constructed the movement index as a quantifiable indicator of a patient’s global amount of movement during a time interval. It is based on variance, which is robust to translation and rotation, and is normalized to account for body size and camera distance. We observed statistically significant and intuitive correlations with GCS and RASS. Patients with the lowest GCS scores had the lowest median movement index, whereas in patients with GCS 15 we observed the highest median movement index. With RASS scores, we noted an inflection point in movement between RASS -1 and 0, corresponding to typical targets for light sedation.

To our knowledge this is the first report demonstrating the use of pose AI in ICU patients. Our results suggest that movement index can be a quantifiable metric of activity and can be used for continuous neuro-telemetry. This addresses important limitations in the current standard of care. Validated exam instruments like GCS and RASS have high interrater reliability but are often inadequate for patients with frequent fluctuations in critical neurological conditions and for monitoring sedation needs. As the GCS and RASS both rely on physical exams by trained providers, delays in obtaining these scores are further exacerbated by staffing limitations.

Video-EEG and BIS have been considered for usage as possible modes of continuous neuromonitoring, but suffer from similar constraints requiring specialized equipment and personnel as well as lacking immediate and intuitive interpretation ^14,15^.

In contrast, pose AI is much more scalable with widely available and low-cost equipment, and is easily and flexibly applicable without special training. Our movement index is highly interpretable as a measurement of variance in movement. A video-based digital health monitoring solution would allow for intuitive review of video footage, pose tracking, and relevant predictions. This technology may reduce the high cost of sedation by optimizing medication selection, titration, and weaning as well as minimizing adverse events. Similar to cardiorespiratory telemetry, pose AI can be easily integrated into the clinical workflow: from a continuous video feed, pose AI will algorithmically provide predictions of neurological status and notify providers when it detects significant deviations from a patient’s baseline movement patterns. Furthermore, variations of the movement index can be developed to recognize other neurological conditions with distinctly abnormal movement patterns, such as seizure and stroke. In 2023, the US National Institutes of Health National Institute of Neurological Disorders and Stroke launched an initiative to expand upon the GCS with a multidimensional framework incorporating blood-based biomarkers, imaging characteristics, and other modifiers. As AI-driven analysis of neurological function continues to evolve, we anticipate that computer vision will play a crucial role in this framework.

## Limitations

There are important limitations to our study that warrant further research. The models were applied out-of-the-box, which does not reflect this technology’s full potential. Targeted pre-training and fine tuning on datasets of real patients in the clinical environment will drastically boost performance. To those points, in our cohort Sapiens is more likely to recognize human figures that are upright rather than lying down, possibly reflecting that it was trained on a non hospitalized population. We also acknowledge that our movement index only accounts for the gross motor aspect of the neurological exam. Future developments in multimodal AI tracking should incorporate other important aspects such as eye opening, facial movement, speech, and cognition. In addition, our video data is collected from patients who underwent video-EEG in a single institution. External validity of our findings needs to be further investigated in broader clinical contexts with multi-institutional data. This study is also vulnerable to biases inherent to the retrospective design. Prospective trials with standardized and optimized video capture and multicenter data collection is a needed next step. Extensive work will be needed to develop a system that effectively integrates this technology into the clinical workflow.

## CONCLUSION

In our study, we externally validated pose AI model performance and derived a movement index that successfully correlated with depth of consciousness as measured by GCS and RASS in NSICU patients. Our findings suggest that pose AI has the potential to serve as a minimally invasive, low-cost, and scalable mode of continuous neuro-monitoring in the NSICU. As neurological conditions account for the highest global disease burden, pose AI may be a flexible solution to address a critical need for neuro-telemetry.

## Data Availability

All data produced in the present study are available upon reasonable request to the authors

