## Supplemental Figure 1, Supplemental Table 1 for "Pose AI prediction of neurological status in the Neuroscience Intensive Care Unit"

**SUPPLEMENTAL MATERIALS**

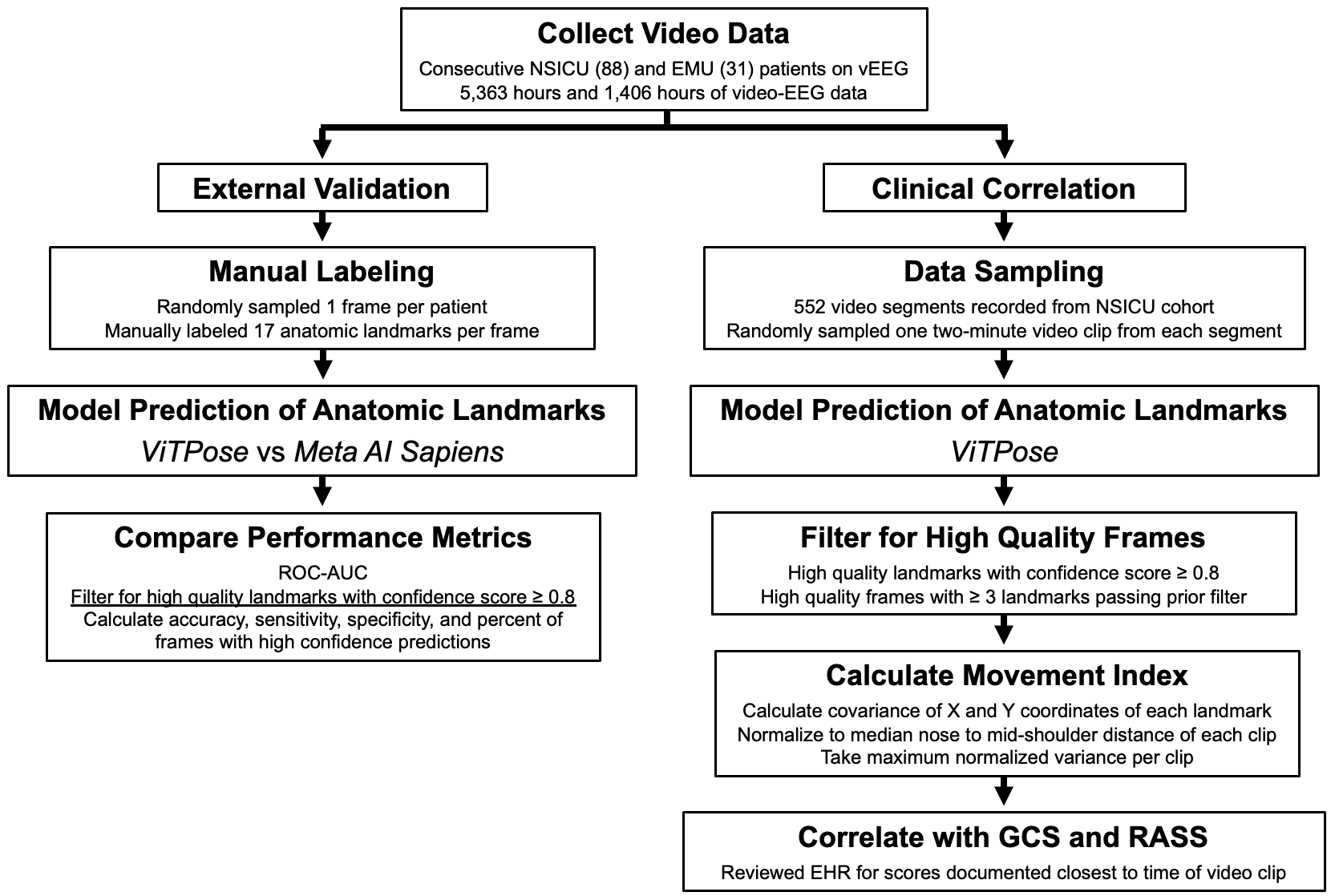

**Supplemental Figure 1. Data processing workflow.** The full data workflow including filters, sample sizes for each group, and evaluation metrics, comporting with TRIPOD guidelines for AI models in healthcare. The left workflow describes our external validation of two leading Pose AI models and the right workflow describes our construction of a robust movement index and correlation with multiple extensively validated states of consciousness.

**Supplemental Table 1. ViTPose median pixel error per anatomic landmark.**

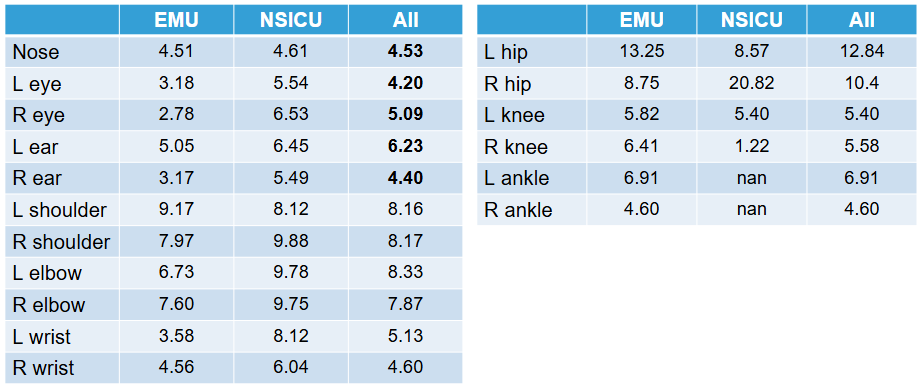
